# Adolescent pregnancy prevalence, maternal characteristics, and perinatal outcomes

**DOI:** 10.1101/2024.08.16.24312096

**Authors:** Paola Toapanta-Pinta, Mercy Dolores Rosero-Quintana, Mayra Soraya Chavez-Espinosa, Cristhian Santiago Vasco-Toapanta, Investigacion en Salud Infantil y Perinatal (ISIP), Santiago Vasco-Morales

## Abstract

**Introduction:** Adolescent pregnancy is a global public health issue with significant implications for maternal and neonatal health, particularly in regions with limited access to healthcare services.

**Objective:** To investigate the prevalence of adolescent pregnancies and perinatal outcomes at the Isidro Ayora Gyneco-Obstetric Hospital during the period 2009-2022.

**Method:** A cross-sectional study was conducted based on records from the Perinatal Information System. Adolescent women and their newborns were included. Multiple pregnancies and cases with incomplete data were excluded. The Chi-square test was applied, and multinomial logistic regression models were used.

**Ethical Approval:** 009-DOC-FCM-2023

**Results:** Out of a total of 26,236 live births, 6,700 (25.53%) were born to adolescent mothers. Many of these mothers were of mestizo ethnicity (94.91%) and had secondary education (80.28%). Multivariate analyses indicated that younger adolescents (<14 years) belonged to minority ethnic groups, substance abuse during pregnancy, underwent episiotomy, and their neonates had a low Apgar score in the first minute of life.

**Conclusions:** Adolescent pregnancy remains a public health issue in Ecuador, associated with adverse neonatal outcomes. It is crucial to implement health policies that address the socioeconomic and cultural determinants of adolescent pregnancy and to conduct prospective studies to better understand the factors involved in these perinatal outcomes.

## INTRODUCTION

According to the World Health Organization (WHO), adolescence is understood to be between 10 and 19 years old.^1^ Studies on social and cognitive development further classify adolescence into three stages: early adolescence, which lasts until the age of 14; middle adolescence, from 14 to 16 years of age; and late adolescence, from 17 to 19.^2,3^

Adolescent pregnancy is a global issue of significant relevance, with profound implications for both maternal and neonatal health. This public health challenge is often linked to socioeconomic and cultural factors that directly affect the quality of life for adolescents and their children. The WHO estimates that 21 million adolescents under the age of 20 become pregnant each year, with the prevalence being higher in low- and middle-income countries. The rate of adolescent pregnancy varies considerably across different regions and socioeconomic contexts, being more common in areas with limited access to sex education, reproductive health services, and contraceptive methods. Between 2015 and 2020, an estimated 62 million babies were born to mothers aged 15 to 19 worldwide. ^4, 5^

In this context, it has been reported that women under 18 years of age face between two and five times the risk of mortality compared to women between 20 and 29 years of age. Over the past few decades, several studies have highlighted that pregnant adolescents face higher risks of obstetric complications and adverse outcomes compared to adult women. These risks include preterm birth, low birth weight, preeclampsia, and higher rates of maternal and infant mortality, including neurodevelopmental disorders. ^6, 7–10^

The health and social implications stemming from teen pregnancy include an increased risk of domestic violence, mental health issues, substance use, sexually transmitted infections, financial stress, and homelessness. Not only do these factors affect the physical and emotional health of pregnant teens, but they can also disrupt their education and personal development, limiting their future opportunities. ^11^

In Ecuador and Latin America, adolescent pregnancy remains a public health problem, requiring multidisciplinary approaches that address both social determinants and risk factors, along with the implementation of effective policies to reduce its prevalence. Considering the aforementioned factors, it is important to understand the incidence, socio-demographic and health characteristics associated with adolescent mothers. ^12^

The objective of this study was to investigate the prevalence of adolescent mothers and perinatal outcomes at the Isidro Ayora Gynecological-Obstetric Hospital between 2009 and 2022.

## METHODOLOGY

*Design and Population.* This was a cross-sectional study based on records from the Perinatal Information System (SIP). The study included adolescent women who had a live newborn and received postpartum surveillance, both mother and newborn, at the Isidro Ayora Gynecological-Obstetric Hospital (HGOIA) from January 2009 to December 2022. This institution is a tertiary-level national reference hospital located in Quito, Ecuador, and is part of the National Public Health System of Ecuador, specializing in the care of teenage pregnancies. Patients with multiple pregnancies and those with incomplete data were excluded from the study. Consecutive sampling was employed.

*Procedure.* Initially, ICD-10 records for Spontaneous Single Birth (O80.0) and Other Assisted Single Births (O83) were identified. The SIP database was subsequently accessed to obtain information on the baseline characteristics of pregnant adolescents, as well as clinical variables related to labor, delivery, and the newborn. This data was filtered and exported to a spreadsheet, where a quality check was performed, and missing data considered lost were identified.

*Variables measured:* Maternal variables included age (categorized as: early adolescence, up to 14 years; middle adolescence, between 14 and 16 years; and late adolescence, from 17 to 19 years and 11 months), educational level, ethnic self-identification, marital status, previous vaginal births and/or cesarean sections, previous abortions, pregnancy planning, alcohol consumption, tobacco and substance abuse during pregnancy, participation in childbirth preparation programs, diagnosis of gestational diabetes mellitus (GDM), development of hypertensive disorders during pregnancy, diagnosis of anemia during pregnancy, diagnosis of urinary tract infection during pregnancy, threat of preterm labor, number of prenatal check-ups, premature rupture of membranes (RPM) or rupture of membranes lasting more than 18 hours, presence of a companion during childbirth, episiotomy, and perineal tears. Regarding newborns (NB), the variables considered included gestational age, birth weight and weight adjusted according to gestational age, Apgar score at the first and fifth minutes, presence of congenital defects, breastfeeding at discharge, and mortality.

Multiple pregnancies were excluded due to the unique characteristics and risks associated with this type of pregnancy, which necessitate analysis in a separate study.

*Analysis:* The data were analyzed with the R programming language (version 4.3.0), “Rcmdr” and “EZR” packages. Qualitative variables were recoded as nominal, dichotomous, or interval. Both absolute and relative frequency were calculated. Baseline variables are present among the group that underwent episiotomy and did not undergo ^13^ this intervention. The crude Odds Ratio (OR) and the adjusted OR (OR a) were estimated with a 95% confidence interval.

Multinomial logistic regression models were used to estimate the OR, adjusting for maternal confounding variables. To select variables for inclusion in the final model, the following steps were performed: (1) selection of variables with the lowest Akaike information criterion (AIC) value in the simple model; (2) model adjustment using the conditional likelihood test; and (3) inclusion of variables with minimum deviance. To address the issue of multiple comparisons, a significance level of 0.01 was used, along with the calculation of the 99% confidence interval (CI) for both crude and adjusted Ors

*Ethical aspects:* This study was approved by the Human Research Ethics Committee of the Central University of Ecuador (CEICH-UCE) code 009-DOC-FCM-2023. As it was a study carried out in an anonymized database, the signing of an informed consent was not required.

## RESULTS

A total of 26 236 newborns were found, of which 6 700 (25.53%) were children of adolescent mothers.

The sociodemographic variables showed that 1.10% (n=74) of the adolescents belonged to the group of early adolescents, while 29.04% (n=1946) corresponded to the group of intermediate adolescents and 69.85% (n=4680) to the group of late adolescents. Regarding ethnic composition, 94.91% (n=6359) were of mestizo ethnicity. Regarding schooling, 80.28% (n=5379) had secondary education. In terms of marital status, 52.93% (n=3546) had a stable partner. In addition, 17.52% (n=1174) had experienced previous pregnancies and 8.12% (n=544) had had previous miscarriages.

Pregnancy variables showed that 23.19% (n=1554) of the cases had planned pregnancy. In addition, 2.46% (n=165) reported alcohol use during pregnancy, while substance abuse was 0.73% (n=49) and tobacco use was 1.37% (n=92). Regarding complications during pregnancy, 0.28% (n=19) of the adolescents had developed gestational diabetes, 18.34% (n=1129) had experienced hypertensive disorders, and 10.46% (n=701) had suffered from anemia. In addition, 41.79% (n=2800) had suffered from urinary tract infection and 16.40% (n=1099) had faced the threat of preterm labor. In terms of prenatal preparation and care, 36.43% (n=2441) of the women received preparation for childbirth, and 43.55% (n=2918) underwent between 5 and 7 prenatal check-ups.

Variables related to childbirth showed that 29.27% (n=1961) of the cases experienced premature rupture of membranes. There was a companion during delivery in 28.81% (n=1930) of the cases. Regarding the method of delivery, 58.43% (n=3915) had vaginal delivery, and of these women, 57.21% (n=2240) underwent episiotomy. In addition, 14.86% (n=582) suffered tears during childbirth.

Variables related to the newborn indicated that 64.90% (n=4348) were born at term. In addition, 52.18% (n=3496) had an adequate birth weight, and 64.91% (n=4349) had an adequate weight for gestational age. 21.90% (n=1407) of the newborns had an Apgar score of less than 7 at the first minute, and 10.42% (n=698) had an Apgar score of less than 7 at the fifth minute after birth. In addition, 11.79% (n=790) had congenital defects. Regarding exclusive breastfeeding at hospital discharge, 94.73% (n=6521) of the newborns were fed in this way. The neonatal mortality rate was 5.27% (n=353).

When comparing the sociodemographic characteristics and pregnancy between the groups of adolescent mothers according to their age, it is evident that in the group of younger adolescents (10–14 years old) there is a higher percentage of belonging to minority ethnic groups. Absence of a stable partner, lack of history of previous pregnancies and therefore miscarriages (Table 1).

**Table 1.**
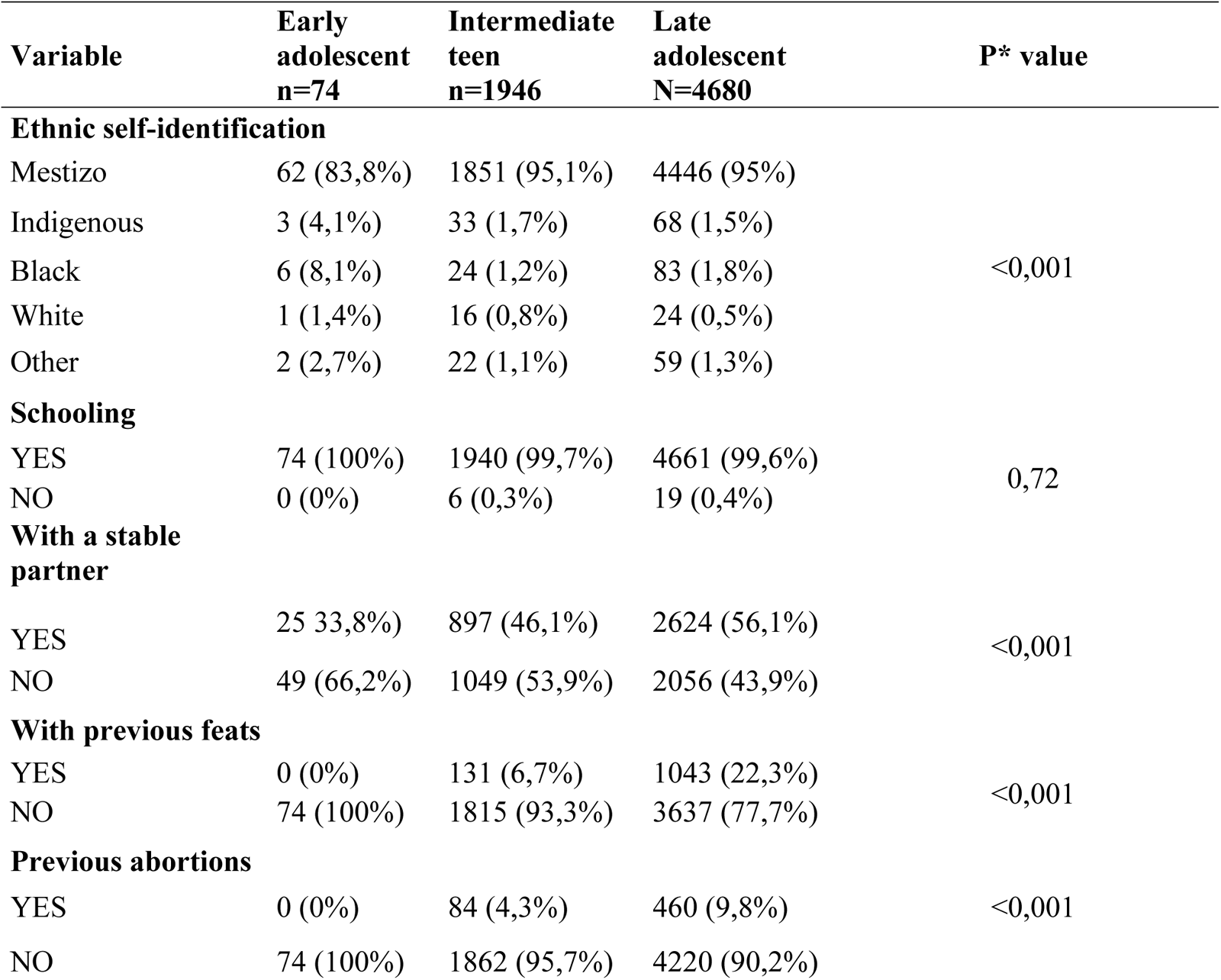
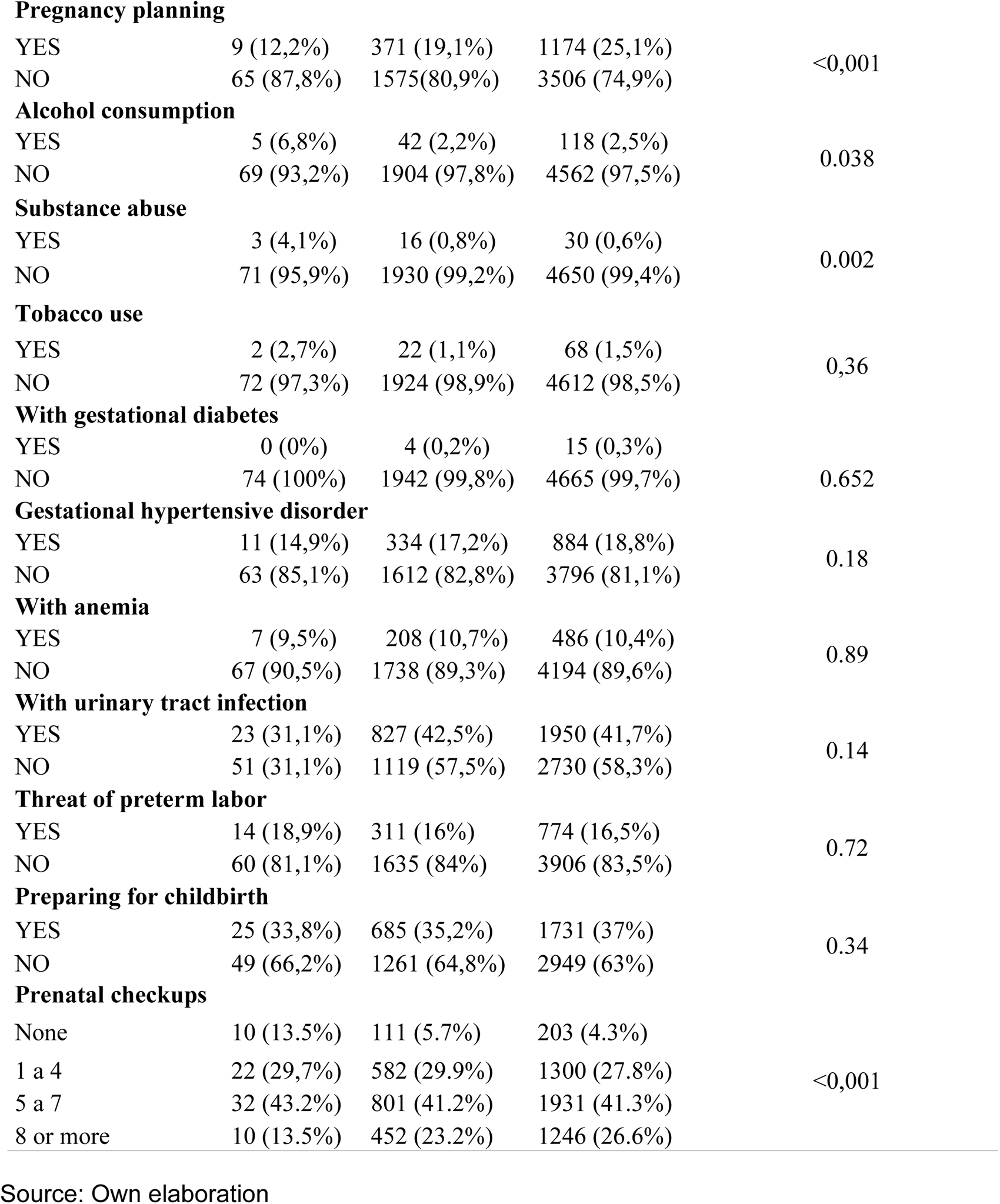
Sociodemographic and pregnancy characteristics, by categories of adolescent mothers, HGOIA, 2009 - 2022.

Late adolescents had a higher percentage of cesarean sections, while early adolescents showed a higher percentage of episiotomy (Table 2).

**Table 2:** Characteristics of childbirth and perinatal outcomes, by categories of adolescent mothers, HGOIA, 2009 - 2022.

| Variable | Early adolescent<br>n=74 | Intermediate teen<br>n=1946 | Late adolescent<br>N=4680 | p* value |
| --- | --- | --- | --- | --- |
| **With premature rupture of membranes** |  |  |  |  |
| YES | 18 (24,3%) | 536 (27,5%) | 1407 (30,1%) | 0.078 |
| NO | 56 (75,7%) | 1410 (72,5%) | 3273 (69,9%) |  |
| **With a companion during childbirth** |  |  |  |  |
| YES | 26 (35,1%) | 572 (29,4%) | 1332 (28,5%) | 0.35 |
| NO | 48(64,9%) | 1374 (70,6%) | 3348 (71,5%) |  |
| **With cesarean section** |  |  |  |  |
| YES | 19 (25,7%) | 733 (37,7%) | 2033 (43,4%) | <0,001 |
| NO | 55 (74,3%) | 1213 (62,3%) | 2647 (56,6%) |  |
| **With episiotomy †** |  |  |  |  |
| YES | 43 (78,2%) | 760 (62,7%) | 1437 (54,3%) | <0,001 |
| NO | 12 (21,8%) | 453 (37,3%) | 1210 (45,7%) |  |
| **With tears** |  |  |  |  |
| YES | 7 (12,7%) | 153 (12,6%) | 422 (15,9%) | 0.023 |
| NO | 48 (87,3%) | 1060 (87,4%) | 2225 (84,1%) |  |
| **Gestational age** |  |  |  |  |
| Preterm | 18 (24,3%) | 661 (34%) | 1455 (31,1%) | 0.004 |
| To the end | 55 (74,3%) | 1240 (63,7%) | 3053 (65,2%) |  |
| Postterm | 1 (1,4%) | 45 (2,3%) | 172 (3,7%) |  |
| **Birth weight** |  |  |  |  |
| Low | 37 (50%) | 920 (47,3%) | 2118 (45,3%) | 0.024 |
| Adequate | 36 (48,6%) | 1004 (51,6%) | 2456 (52,5%) |  |
| Elevated | 1 (1,4%) | 22 (1,1%) | 106 (2,3%) |  |
| **Weight-for-gestational age** |  |  |  |  |
| Small | 18 (24,3%) | 661 (34%) | 1454 (31,1%) | 0.004 |
| Adequate | 55 (74,3%) | 1240 (63,7%) | 3054 (65,3%) |  |
| Big | 1 (1,4%) | 45 (2,3%) | 172 (3,7%) |  |
| **Apgar less than 7 at the first minute** |  |  |  |  |
| YES | 25 (33,8%) | 425 (21,8%) | 957 (20,4%) | 0.011 |
| YES | 11 (14,9%) | 194 (10%) | 493 (10,5%) |  |
| **Apgar less than 7 at the fifth minute** |  |  |  | 0.35 |
| YES | 25 (33,8%) | 425 (21.8%) | 957 (20.4%) |  |
| NO | 49 (66,2%) | 1521 (78.2%) | 3723 (79.6%) |  |
| **With birth defects** |  |  |  |  |
| SI | 10 (13,5%) | 237 (12.2%) | 543 (11.6%) |  |
| NO | 64 (86.5%) | 1709 (87.8%) | 4137 (88.4%) | 0.7217 |
| **Breastfed at discharge** |  |  |  |  |
| YES | 73 (98,6%) | 1896 (97,4%) | 4552 (97,3%) |  |
| NO | 1 (1,4%) | 50 (2,6%) | 128 (2,7%) | 0.7237 |
| **Live neonate** |  |  |  |  |
| YES | 67 (90,5%) | 1832 (94,1%) | 4448 (95%) |  |
| NO | 7 (9,5%) | 114 (5,9%) | 232 (4,9%) | 0.08768 |

\*Chi 2, † n=2240
Source: Own elaboration

In the simple multinomial logistic regression analysis, the category of early adolescent pregnancy is used as a reference point. It is observed that middle-aged and late adolescents have a lower risk of belonging to minority ethnicities, alcohol consumption, substance abuse, episiotomy and having an Apgar score of less than 7 per minute. It is also shown that these groups have a higher probability of having a stable partner and a greater number of prenatal check-ups.

In addition, late-aged adolescents are more likely to have a planned pregnancy, to have a cesarean delivery, and a lower likelihood of tobacco use.

The variables that produced infinite confidence intervals in the simple multinomial logistic regression analysis are not presented.

Multiple multinomial logistic regression analysis confirmed that middle- and late-adolescent mother groups are less likely to belong to an ethnic minority and to undergo an episiotomy compared to younger adolescent girls. In addition, late adolescents are less likely to engage in substance abuse, and their babies are less likely to have an Apgar score of less than 7 in the first minute of life (Table 3).

**Table 3.** Variables associated with age categories in adolescent pregnancy HGOIA, 2009 - 2022.

| Variables | Simple multinomial logistic regression |  | Multiple multinomial logistic regression |  |
| --- | --- | --- | --- | --- |
|  | Middle adolescence<br>OR bruto (IC 95%) | Late adolescence<br>OR bruto (IC 95%) | Middle adolescence<br>OR ajustado (IC 95%) | Late adolescence<br>OR ajustado (IC 95%) |
| **Minority ethnicity** | 0,26 (0,13; 0,50) | 0,27 (0,13 ;0,50) | 0,25 (0,11; 0,57) | 0,24 (0,11; 0,54) |
| **With a stable partner** | 1,68 (1,03; 2,74) | 2,50 (1,54; 4,06) | 1,27 (0,71; 2,25) | 1,69 (0,95; 2,99) |
| **With pregnancy planning** | 1,70 (0,84; 3,45) | 2,42 (1,2; 4,87) | 1,44 (0,65; 3,18) | 1,74 (0,79; 3,83) |
| **With alcohol consumption** | 0,30 (0,11; 0,79) | 0,35 (0,14; 0,90) | 0,43 (0,11; 1,59) | 0,49 (0,13; 1,80) |
| **Substance abuse** | 0,19 (0,05; 0,68) | 0,15 (0,045; 0,51) | 0,25 (0,05; 1,23) | 0,12 (0,02; 0,58) |
| **With tobacco use** | 0,19 (0,05; 0,68) | 0,15 (0,04; 0,51) | 1,09 (0,17; 6,83) | 1,38 (0,22; 8,41) |
| **Prenatal checkups** | 1,13 (1,03; 1,24) | 1,17 (1,07; 1,29) | 1,15 (0,82; 1,60) | 1,24 (0,88; 1,73) |
| **With cesarean section** | 2,22 (1,03; 2,97) | 2,22 (1,32; 3,76) | 1 (1; 1) | 1 (1; 1) |
| **With episiotomy** | 0,46 (0,24; 0,89) | 0,33 (0,17; 0,63) | 0,29 (0,21; 0,82) | 0,29 (0,15; 0,57) |
| **With Apgar less than 7 per minute** | 0,54 (0,33; 0,89) | 0,50 (0,31; 0,82) | 0,59 (0,32; 1,06) | 0,53 (0,30; 0,96) |

## DISCUSSION

The present study found that 25.53% of newborns in the HGOIA are children of adolescent mothers. This figure is like that previously reported in Ecuador in 2016, where 24,794 adolescent births were registered, representing 26% of the total births, of which 7% corresponded to girls between 10 and 14 years old. ^14^ While data from the Ecuadorian Institute of Statistics and Census (INEC) shows that, in the 1990s, the birth rate to mothers aged 10 to 14 was 0.11%, this figure increased to 0.23% in 2021. In the case of mothers aged 15 to 19, the rate was 7.36% in the 1990s, falling to 4.73% in 2022. On the other hand, according to recent epidemiological studies, the proportion of births to mothers under 18 years of age in Ecuador (9.9%) is still higher than in countries such as the United States (1.5%) or Canada (0.9%). The high prevalence observed in the present study could be partially explained by the role of the HGOIA as a national reference hospital, which has a specialized service in the care of pregnant adolescents.^15, 16^

Globally, the rate of teenage pregnancies has decreased, going from an average of 9.00% among women aged 15 to 19 in the 1960s to 4.25% in 2021, according to World Bank data. However, regional differences persist in Europe, Central Asia and North America, rates are less than 2.00%, while in Latin America they vary between 1.81% in Chile and 8.56% in Nicaragua. In sub-Saharan Africa, rates range from 2.46 in Mauritius to 17% in Niger. ^17^

In contrast, epidemiological studies reveal more specific variations, for example, in the Eastern Mediterranean region, a systematic review estimated a prevalence of 9%, while in Africa, the combined prevalence reached 18.8%, reaching 19.3% (95% CI: 16.9, 21.6) in the sub-Saharan African region. ^18^ The highest prevalence was recorded in East Africa (21.5 per cent) and the lowest in North Africa (9.2 per cent). ^19^ If the rates are analyzed by country, we have, for example, Tanzania, a cross-sectional community study reported a rate of 29%. ^20^ In Sierra Leone, the prevalence of teenage pregnancies was 22.1 per cent, according to the Demographic and Health Survey. In Uganda, a study conducted in a referral hospital found a prevalence of 20.6% (95% CI: 17.0% - 24.7%). ^21, 22^ In Ethiopia, a cross-sectional study in schoolgirls reported a prevalence of 14.6% (95% CI: 11.9% - 17.7%), while another study found that 12.2% (95% CI: 9.5% - 14.9%) of pregnant women who attended a health institution were adolescents. ^23, 24^

The differences between epidemiological studies and data from the World Bank and INEC are since the latter are aggregated and standardized for international comparisons, while epidemiological studies focus on specific contexts and may reflect local variations. ^25, 26^

In short, in Latin America, the fertility rate has decreased significantly in women aged 25 to 29, but the reduction has been slower in adolescents aged 15 to 19 years. The region shows the slowest decline in adolescent fertility globally (15%) and is the only one where fertility has increased in children under 15 years of age. This situation is more common in low- and middle-income countries, where it is associated with worse obstetric outcomes, such as preeclampsia, low birth weight, and increased risk of infections during pregnancy. ^19, 25, 27^

### Ethnic self-identification

This study found that middle and late adolescents were less likely to belong to minority ethnicities relative to early adolescents. In this regard, Mann et al. (2020) observed that in Australia, adolescent pregnancy rates are increasing, with a more pronounced trend among Aboriginal and Torres Strait Islander adolescents compared to non-indigenous adolescents. However, no differences are noted between the different age groups.^11^

Research shows that teen pregnancy rates disproportionately affect African, Caribbean, and Black teens in the U.S., while the lowest rates in 2022 were for Asians and whites, at 0.19% and 9.1%, respectively. In addition to age and ethnicity, there are other intervening factors such as socioeconomic and cultural factors, along with limited access to reproductive health services, are key determinants of these racial disparities. ^28,29^

### Substance abuse

The findings of our study, which reveal a high risk of consumption of psychoactive substances during pregnancy in adolescent mothers, particularly in younger mothers, are consistent with the existing literature. In this sense, Wong et al. (2020), reported a significantly higher prevalence of tobacco, marijuana, and alcohol use among young mothers compared to their adult counterparts (P < 0.001), which can be associated with poor outcomes in the newborn such as low Apgar. ^30^

### Episiotomy

In the group examined in the present study, it was found that 57.21% of adolescent mothers who had normal delivery underwent episiotomy, with a higher frequency in early adolescents who showed a significant difference in relation to middle and late adolescents. In a study carried out at the HGOIA, in 11862 patients, a positive association was found with this practice and maternal age less than 20 years. In the study by Abebe et al. (2020), it was observed that adolescent pregnancy was related to episiotomy (AOR: 2.01; 95% CI, 1.25-3.39). Similarly, Okumura et al. (2014) also indicated that adolescent girls have a higher risk of episiotomy (OR=1.34; 95%CI=1.29-1.40). However, current guidelines advise against routine episiotomy.^31, 32, 33, 31^

### Low Apgar

In this study, 21.9% of newborns had an Apgar score of less than 7 in the first minute of life, being more common among children of early adolescents. The RW confirmed that late teens are less likely to have a low first-minute Apgar score compared to early teens. In a previous study carried out in this same population, which compared adult and adolescent women, no significant differences were found in the Apgar score; however, in this study, adolescents were not stratified according to their age, which could explain the absence of differences. ^34^

In research by Ogawa et al. (2019), adolescent pregnancy was found to be associated with a higher incidence of low Apgar scores (RR 1.41, 95% CI 1.15–1.73) compared to women aged 20–24 years. Similarly, Egbe et al. (2019) observed that the highest proportion of infants with low Apgar scores (<7) at one minute of life was among children of adolescent mothers, with 7.1% versus 3.1% in other age groups (p = 0.01). Radu et al. (2022), reported that 23.4% of newborns born to adolescent mothers had Apgar scores below seven at the first minute of life. In addition, Diabelková et al. (2023) found that children of adolescent mothers had significantly lower Apgar scores in the first minute (p = 0.003). It is important to note that, in addition to maternal age, other factors such as the use of psychoactive substances, including tobacco, marijuana, and alcohol during pregnancy, can negatively influence neonatal outcomes. ^35, 36, 37, 38, 30^

The present study has inherent limitations in its design and the use of secondary data from a single hospital, which may have resulted in the omission of relevant information. Despite this, the findings highlight the need for future prospective research that also analyzes factors associated with fathers.

The present work presents strengths, such as the historical evaluation of this problem, due to the size of the sample and the period studied, it provides comparable data at the national and international level.

## CONCLUSIONS

This study revealed that 25.53% of newborns in the HGOIA are children of adolescent mothers, underscoring the persistence of adolescent pregnancy as a public health problem in Ecuador. The results show that adolescent pregnancies are significantly associated with adverse neonatal outcomes, including low Apgar scores, an increased risk of episiotomy, and an increase in the use of psychoactive substances during pregnancy, especially among younger adolescents. In addition, ethnic self-identification and socioeconomic conditions continue to be determining factors influencing adolescent pregnancy rates

## RECOMMENDATIONS

Prospective, multicenter studies are required that include a broader and more complete analysis of the characteristics of adolescent mothers and the factors that influence perinatal outcomes. Research and development of programs that include adolescent boys should be encouraged, since their role in the prevention of adolescent pregnancy is an aspect that has been underestimated in the existing literature.

## Data Availability

All data produced in the present work are contained in the manuscript

https://data.mendeley.com/datasets/y4s9khhs5k/2

